# Prevalence and Factors Associated with Renal Disease Among HIV Infected Children on Antiretroviral Therapy Attending Joint Clinical Research Center, Lubowa, Kampala

**DOI:** 10.1101/2023.02.09.23285685

**Authors:** Lynn Kolosi Bagoloire, Ansiima Sheilla, Joan Kalyango, Henry Mugerwa, Kaggwa Bruhan, Peter Waiswa, Nicolette Nabukeera Barungi, Victor Musiime

**Affiliations:** Clinical Epidemiology Unit, School of Medicine, Makerere University College of Health Sciences, P.O Box 7062, Kampala, Uganda; Department of Pediatrics and Child Health, School of Medicine, Makerere University, College of Health Sciences, P.O Box 7062, Kampala, Uganda; Department of Pharmacy, Makerere University, College of Health Sciences, P.O Box 7062, Kampala, Uganda; Baylor College of Medicine Children’s Foundation, Block 5 Mulago Hospital, P.O Box 72052, Kampala, Uganda; Joint Clinical Research Center, Lubowa, P.O Box 10005, Kampala, Uganda; School of Public Health, Makerere University, P.O Box 7062, Kampala, Uganda

## Abstract

**Introduction:** Renal disease is asymptomatic in its early stages but on progression into advanced stage is expensive to manage requiring renal replacement therapy. Monitoring of children’s renal functionality is not routinely done but only on a doctor’s request. However, there is currently no published study on renal disease prevalence among children living with HIV in Uganda and the associated factors.

**Objective:** To determine the prevalence of renal disease and its associated factors among HIV positive children on ART at the Joint Clinical Research Center (JCRC) Pediatric clinic.

**Methods:** We conducted a cross sectional study from JCRC Pediatrics Clinic. Children attending the clinic between January and August, 2020 were recruited using consecutive sampling. Information on age, sex, z sore for age, Blood pressure, route of transmission, clinical stage of the disease, ART regimen, duration on ART, non-ART drugs, Viral load and comorbidities was obtained. Blood and urine samples were also collected from the participants and sent to the laboratory for analysis. Renal disease was diagnosed using either proteinuria on urine dipstick or reduced estimated Glomerular Filtration Rate, (eGFR).

**Results:** The study participants were 261, of which 51% were female and 49% male, with a median age of 12 years. The prevalence of renal diseases was 21.5% as proteinuria and 9.6% with low eGFR. Factors associated with proteinuria were; age (p-value =0.01), clinical stage of the disease (p=0.035) and diastolic blood pressure (DBP, p = 0.001) and with low eGFR were; DBP (p < 0.001) and clinical stage of disease (p < 0.001).

**Conclusion:** The prevalence of proteinuria was high (21.5%) and that of low eGFR was (9.1%). Early screening for renal disease when still asymptomatic as a preventive strategy for renal disease should be considered, especially using affordable yet effective methods like urinalysis. This is because all the participants were asymptomatic at the time of enrollment.

## Introduction

Approximately 35 million people worldwide are living with HIV according to the United Nations Program on HIV/AIDS (UNAIDS) and more than 2 million new infections are registered annually (UNAIDS 2016). About 1.8 million children under the age of 15 years are living with HIV globally according to the 2020 UN World AIDs day Report, 90% of these are in sub Saharan Africa (SSA) (UNAIDS 2016). In Uganda, 90,000 children between 0 and 14 years are estimated to be living with HIV, according to a Uganda Population Based HIV Impact Assessment conducted in 2016 to 2017 (Uganda 2016). Successful clinical outcomes on HIV management with antiretroviral therapy (ART), have improved patients’ survival, thereby paving way for long term diseases like renal complications (Ray, Xu et al. 2004). A systematic review conducted by Ekrikpo et al, reported the prevalence of renal disease among HIV adults at 4.8% globally, and Africa’s prevalence at 7.9% (Ekrikpo, Kengne et al. 2018).

Renal disease could arise from HIV infection; starting from the local HIV infection of the kidney, involving viral infection of the tubular and glomerular epithelial cells causing the collapse of the glomerular basement membranes accompanied by hypertrophy and hyperplasia of the overlying glomerular epithelial cells which may fill the urinary space forming pseudo crescents (Swanepoel, Atta et al. 2018).

Different studies have reported ART as a contributing factor to renal complications among HIV patients (ONLIN 2013). Lifelong exposure of HIV positive patients to ART predisposes them to acute and chronic renal disease (Swanepoel, Atta et al. 2018). There have been reports on renal injury from (Tenofovir Disoproxil Fumarate) TDF-based regimen when combined with protease inhibitors (PIs) (Brennan, Evans et al. 2011, Bygrave, Kranzer et al. 2011). The occurrence of renal disease among the HIV-infected population may also be increased by drug-induced renal toxicity, opportunistic infections, and other comorbidities like hypertension, diabetes, and viral infections like hepatitis (Msango, Downs et al. 2011).

Some of the medicines used in HIV management like rifampicin, cotrimoxazole, amphotericin B and antibiotics like aminoglycosides for prophylaxis against opportunistic infections in children and adults have been associated with renal disease (Kalyesubula, Wearne et al. 2014). Malnutrition which is common among children under 5 years in the Ugandan setting, is associated with renal disease (Foster and Leonard 2004). The prevalence of acute malnutrition (wasting) among children under 5 years is at 4% in Uganda (Dutta, Barker et al. 2015). Long-term use of ART, drug toxicity and chronic viral infections have resulted in an increase in the frequency of kidney disease in HIV-infected individuals (Trullàs, Barril et al. 2008). Children are likely to have longer exposure not only to ART but also other medicines and this may predispose them to renal disease.

In Sub-Saharan Africa, a few studies have described the extent of renal disease in HIV-infected children (Iduoriyekemwen, Sadoh et al. 2013) with scattered literature from different African countries. A systematic review conducted in sub Saharan Africa among children revealed a prevalence of 32.5% (Kayange, Smart et al. 2015). Some of the factors associated with renal disease among HIV infected children include; age, WHO clinical staging of the disease, ART regimen and duration on ART (Dondo, Mujuru et al. 2013, Iduoriyekemwen, Sadoh et al. 2013).

Currently, there is limited published data, if any, on renal disease prevalence among HIV infected children on ART in Uganda. This study aimed at identifying the prevalence of renal disease among HIV infected children on ART and the factors associated with renal disease among this population.

## Materials and methods

### Study setting

This study was conducted from the pediatrics clinic at Joint Clinical Research Center which is headquartered in Lubowa along Entebbe Road, less than 9km away from Kampala city. The research center was founded in 1990 and is a collaborative effort by three Ugandan ministries to address the challenges posed by HIV/AIDS and related infections, these include; the Ministry of Health, the Ministry of Education and the Ministry of Defense. The facility receives patients from Wakiso, Kampala, Mityana, Mukono, other parts of central Uganda and on few occasions, from as far as South Sudan. At the time of the study, there were 1,095 children enrolled on ART, 695 children were on 1^st^ line ART, 333 on 2^nd^ line, 40 on 3^rd^ line and 27 unspecified regimens.

### Study design

A cross sectional study was conducted to determine the prevalence of renal disease among HIV infected children on ART and the associated factors from February, 2020 to August, 2020.

### Study population

HIV positive children on ART who were below 18 years, attending JCRC Pediatric clinic and visited between February and August, 2020 and met the eligibility criteria.

### Sample size and sampling

Sample size was calculated using the Kish Leslie formula (Kish 1995) for estimation of sample size in single groups for cross sectional studies. For the determination of factors associated with renal disease, sample size sample size calculation for two proportions was used. The exposure factor was HIV positive children on ART. After adjusting for 10% refusal to participate, 261 participants were enrolled into the study.

### Inclusion Criteria

HIV positive children attending JCRC and are on ART, between 3 and 17 years, had come for review between February and August, 2020, their parents/guardians had given informed consent and those above 8 years who assented.

### Exclusion Criteria

Participants whose information like age, did not correspond with the information in their clinic files and those who were febrile with preexisting symptomatic urinary tract infections

### Study Variables

The socio-demographic and patient related factors from literature predisposing to renal disease were age, sex, z score for age and blood pressure, as well as route of transmission (ONLIN 2013) (Foster and Leonard 2004) (Dutta, Barker et al. 2015) (Dondo, Mujuru et al. 2013, Iduoriyekemwen, Sadoh et al. 2013). Data on socio-demographics was obtained by administering a questionnaire.

The predisposing patient clinical characteristics were; WHO clinical stage of the disease, ART regimen, duration on ART, co-administered drugs, viral load and comorbidities (Brennan, Evans et al. 2011, Bygrave, Kranzer et al. 2011, Swanepoel, Atta et al. 2018) (Msango, Downs et al. 2011) (Dondo, Mujuru et al. 2013, Iduoriyekemwen, Sadoh et al. 2013). Data on predisposing factors was obtained by review of patient medical records.

### Data Management

Data was double entered by two data entry assistants using Epidata version 3.1. The filled and filed data abstraction forms were stored under lock and key for reference purposes, and also to ensure safe storage and confidentiality. Data was exported into STATA version 14.0 for further cleaning and analysis.

### Data Analysis

Continuous variables following a normal distribution were summarized using means and standard deviations, and those not normally distributed were summarized using medians. Categorical variables were summarized using proportions and percentages.

The prevalence of renal disease was summarized as the number of children having renal disease (either reduced eGFR or proteinuria on urine dipstick) divided by the total number of children in the study and the proportion was multiplied by 100%.

The proportions along with their 95% confidence intervals for age, sex, blood pressure, route of transmission, duration on ART, ART regimen, non-ART regimen, viral load and WHO clinical staging of HIV were also determined.

Logistic regression was used to assess the association between prevalence of renal disease and the independent variables.

Bivariate analysis was performed for each of the independent variables to determine whether they were independently associated with renal disease using odds ratios (OR) and p values. Factors with p value ≤0.2 were considered for multivariate analysis. In multivariate analysis, step wise backward method was used to identify independent variables with p value ≤0.05, and then assessing for interaction by comparing the -2 Log Likelihoods of the reduced and full models. Confounding was assessed for basing on a difference ≥ 10% between the crude and adjusted prevalence ratios of the variables.

### Ethical Considerations

Prior to commencement of the study, permission to conduct the study was sought from the Makerere University Clinical Epidemiological Unit and ethical approval for this study, was obtained from the Makerere University School of Medicine Research Ethics Committee (SOMREC) and finally administrative approval from the Joint Clinical Research Center which was the study site. A written informed consent was obtained from parents/guardians on behalf of the study participants below 8 years and assent obtained from children between 8 and 17 years of age. The children who tested positive with renal disease were forwarded to the JCRC medical team for further clinical attention.

## Results

### Description of Study Participants

Majority were female (50.6%), all the participants had acquired HIV through vertical transmission, 260 of them were taking cotrimoxazole as prophylaxis for opportunistic infections and only 1 was taking isoniazid, none had tuberculosis. Majority of the participants were underweight (71.3%), majority (76.6%) had been on ART for more than 5 years, 76.8% had a low viral load below 10,000 copies/ml, majority (47.9%) were in WHO clinical stage II and majority (36.8%) were on abacavir based regimen, as described in Table 1 below, which is a summary of characteristics of the participants.

**Table 1.**
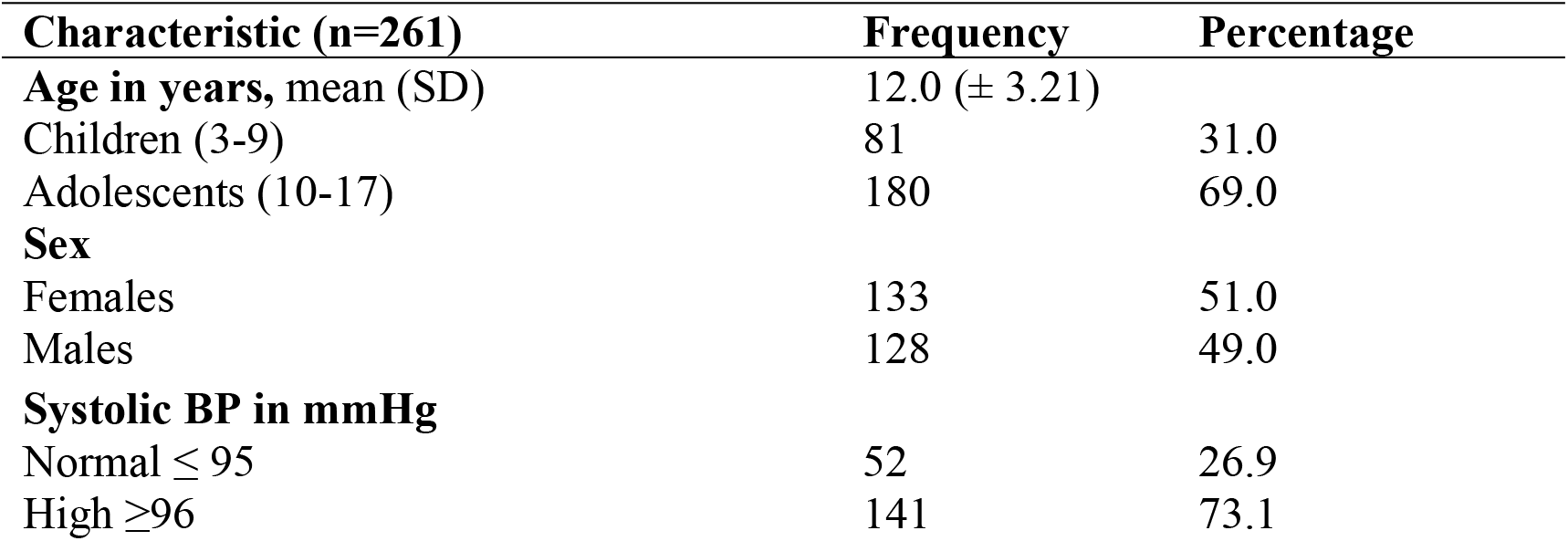
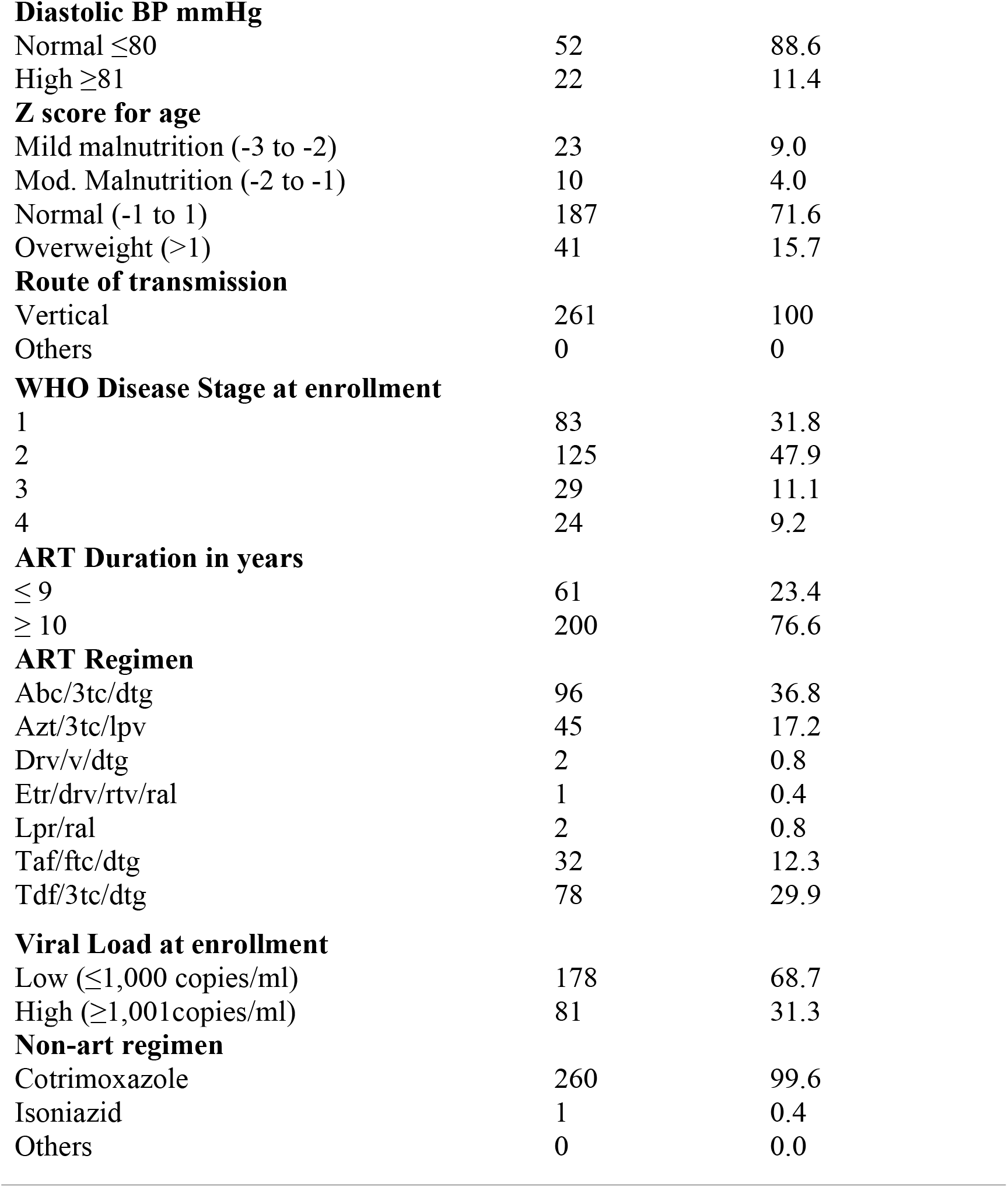
Demographic and Clinical Characteristics of 261 HIV-infected Children Attending JCRC Pediatric Clinic from February, 2020 to August, 2020.

### Prevalence of renal disease

Of the 261 participants studied, the overall prevalence of renal disease was 31.1%; 21.5% (95% CI: 73.0 – 83.0%) had renal disease (proteinuria) on urinalysis and 9.6% (95% CI: 7.0 – 14.0%) had renal disease (low eGFR); 15.4% (95% CI: 50.6 – 76.0) of the children who had a normal eGFR also had proteinuria and 80% (95% CI: 24.0 – 49.4) of those with a low eGFR had proteinuria as well. The prevalence of proteinuria was higher among the females (59% 95% CI 45.0 – 71.0%) as compared to the males. Below are tables 2 and 3, describing the prevalence of renal disease, that is proteinuria and low eGFR, respectively by; age group, sex, blood pressure category, ART duration, viral load, z-score for age, ART regimen, WHO clinical stage of disease, route of transmission and non-ART regimen.

**Table 2.**
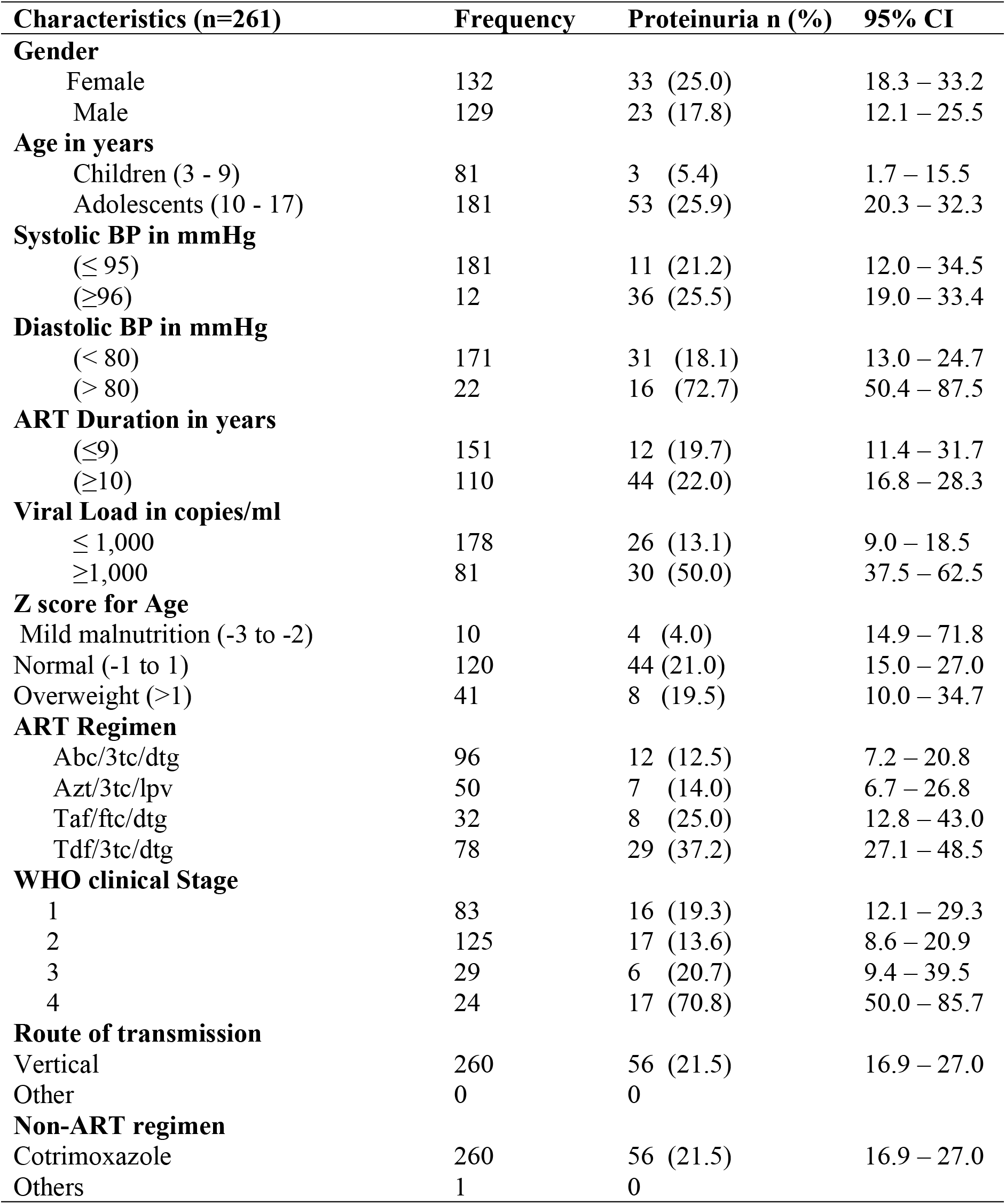
Prevalence of renal disease (Proteinuria) among 261 HIV Positive Children on ART Attending JCRC Pediatrics Clinic from February, 2020 to August, 2020.

**Table 3.**
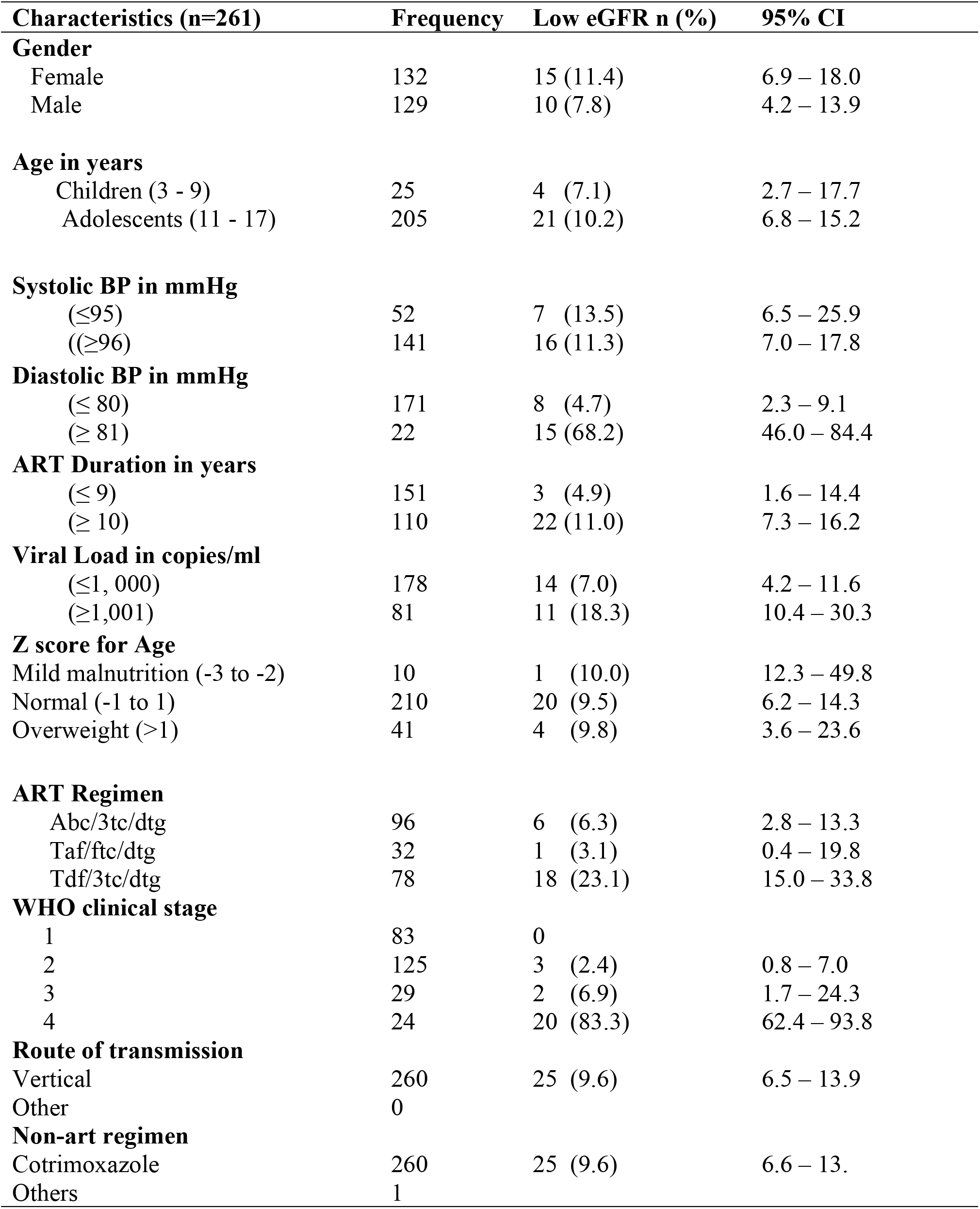
Prevalence of renal disease (low eGFR) among 261 HIV Positive Children on ART Attending JCRC Pediatrics Clinic from February, 2020 to August, 2020.

### Factors Associated with Renal Disease among HIV Positive Children on ART

Analyses of factors associated with viral renal disease among HIV positive children on ART attending JCRC Pediatrics Clinic receiving ART are summarized in tables 4 and 5. On bivariate analysis, the independent variables that were significantly associated with renal disease (proteinuria) were age (OR= 6.2, p value = <0.003), SBP (OR= 1.7, p value = 0.084), DBP (OR = 12.04 p value = <0.001), Viral load (OR= 5.1, p value = 0.001), z score for age on overweight (OR = 7.4, p value = 0.013) and sex (OR =0.65, p value = <0.16). Results of bivariate analyses of factors associated with renal disease are summarised in table 3.

**Table 4.** Bivariate Analysis of Factors Associated with Proteinuria Among HIV-infected Children on ART, 3-17 years old (n=261)

| Variable | Proteinuria<br>n (%) | OR | 95% CI | P-value |
| --- | --- | --- | --- | --- |
| **Age in years** |  |  |  |  |
| Children (3 – 10) | 3 (12.0) | 1 | 1 |  |
| Adolescents (11 - 17) | 53 (25.9) | 6.2 | 1.85 – 22.47 | 0.003 |
| **Gender** |  |  |  |  |
| Female | 4 (4.9) | 1 | 1 |  |
| Male | 52 (28.9) | 0.7 | 0.35 – 1.18 | 0.16 |
| **Systolic BP in mmHg** |  |  |  |  |
| ≤ 95 | 20 (16.7) | 1 | 1 |  |
| ≥ 96 | 36 (10.8) | 1.7 | 0.93 – 3.16 | 0.084 |
| **Diastolic BP in mmHg** |  |  |  |  |
| ≤ 80 | 40 (23.4) | 1 | 1 |  |
| ≥ 81 | 16 (72.7) | 12.0 | 4.36 – 33.26 | <0.001 |
| **ART Duration in years** |  |  |  |  |
| ≤ 9 | 28 (18.5) | 1 | 1 |  |
| ≥ 10 | 28 (45.5) | 1.6 | 0.68 – 3.55 | 0.30 |
| **Viral load in copies/ml** |  |  |  |  |
| ≤ 1,000 | 22 (39.3) | 1 | 1 |  |
| ≥ 1,001 | 34 (42.0) | 6.7 | 2.74 – 9.61 | <0.001 |
| **Z score for age** |  |  |  |  |
| Mild malnutrition (-3 to -2) | 2 (8.7) | 1 | 1 |  |
| Normal (-1 to 1) | 37 (19.8) | 2.6 | 0.58 – 11.54 | 0.212 |
| Overweight (>1) | 17 (41.5) | 7.44 | 1.54 - 36.03 | 0.013 |
| **Drug** |  |  |  |  |
| Abc/3tc/dtg | 12 (12.5) | 1 | 1 |  |
| Azt/3tc/lpv | 7 (14.0) | 1.1 | 0.42 – 3.10 | 0.80 |
| Taf/ftc/dtg | 8 (25.0) | 2.3 | 0.86 – 6.36 | 0.10 |
| Tdf/3tc/dtg | 29 (37.2) | 4.1 | 1.94 – 8.85 | <0.001 |
| **Stage** |  |  |  |  |
| 1 | 16 (19.3) | 1 | 1 | 1 |
| 2 | 17 (13.6) | 0.7 | 0.31 – 1.39 | 0.275 |
| 3 | 6 (20.7) | 1.1 | 0.38 – 3.12 | 0.869 |
| 4 | 17 (70.8) | 10.2 | 3.61 – 28.64 | <0.001 |

**Table 5.**
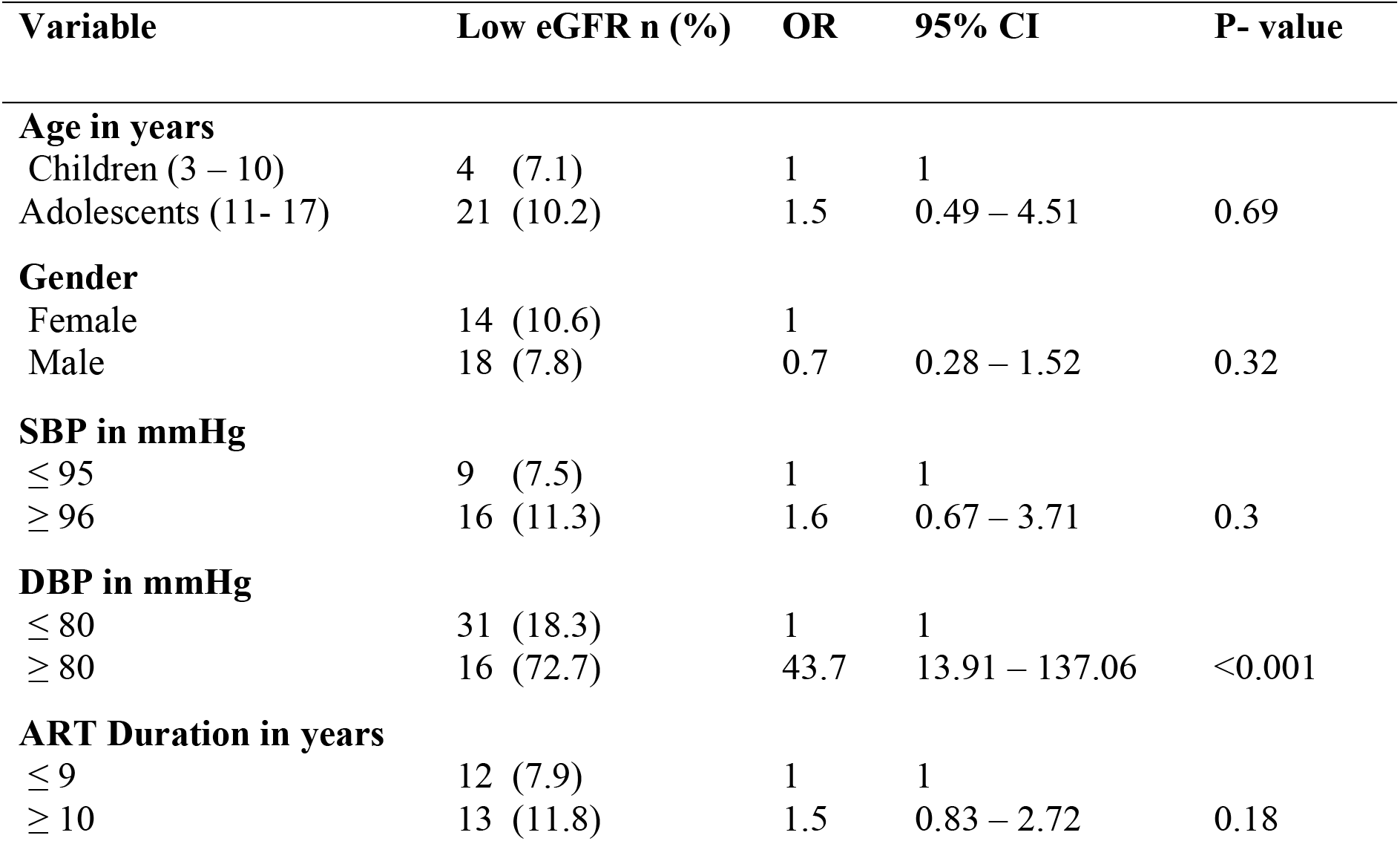
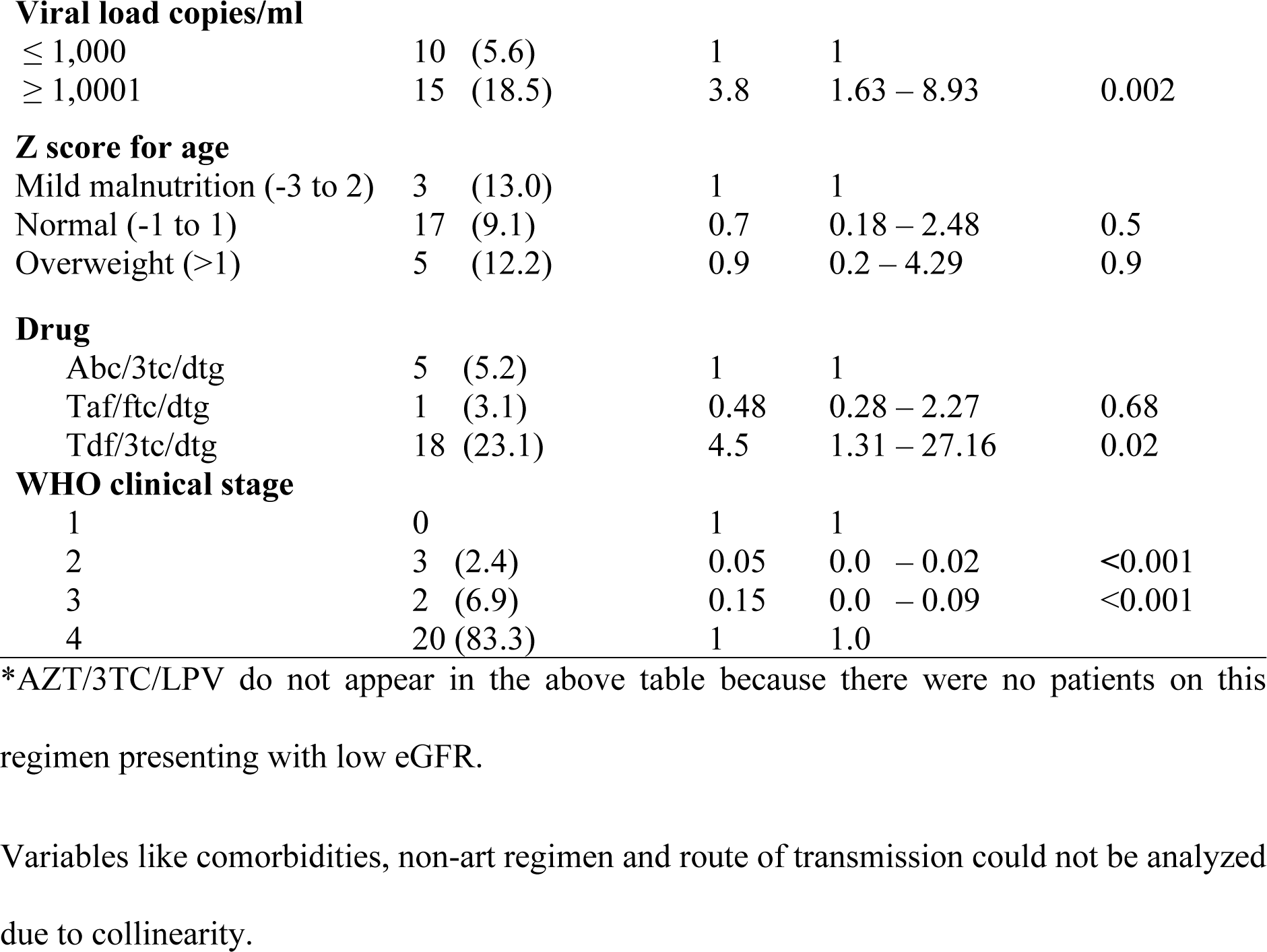
Bivariate analysis of factors associated with renal disease (Low eGFR) among HIV positive children on ART attending JCRC Peadiatrics Clinic.

**Table 6.** Multivariate analysis of factors Associated with Proteinuria Among HIV-infected Children on ART, 3-17 years old (n=261)

| Variable | OR | 95% CI | P-value |
| --- | --- | --- | --- |
| **Age in years** |  |  |  |
| Children (3 – 9) | 1 | 1 |  |
| Adolescents (10 – 17) | 11.8 | 2.79 – 45.71 | 0.001 |
| **Viral load copies/ml** |  |  |  |
| ≤ 1,000 | 1 | 1 |  |
| ≥ 1,000 | 7.0 | 3.29 – 14.96 | < 0.001 |
| **DBP in mmHg** |  |  |  |
| ≤ 80 | 1 | 1 |  |
| ≥ 81 | 6.5 | 1.76 – 23.88 | 0.005 |
| **WHO Disease stage** |  |  |  |
| 1 | 1 | 1 |  |
| 2 | 0.6 | 0.25 – 1.36 | 0.22 |
| 3 | 1.1 | 0.32 – 3.74 | 0.88 |
| 4 | 4.4 | 1.11 – 17.57 | 0.035 |

**Table 7.** Multivariate analysis of Factors Associated with a Low eGFR Among HIV-infected Children on ART, 3-17 years old (n=261)

| Variable | OR | 95% CI | P-value |
| --- | --- | --- | --- |
| **DBP in mmHg** |  |  |  |
| < 80 | 1 | 1 |  |
| > 81 | 40.89 | 12.44 – 134 | <0.001 |

**Table 8.**
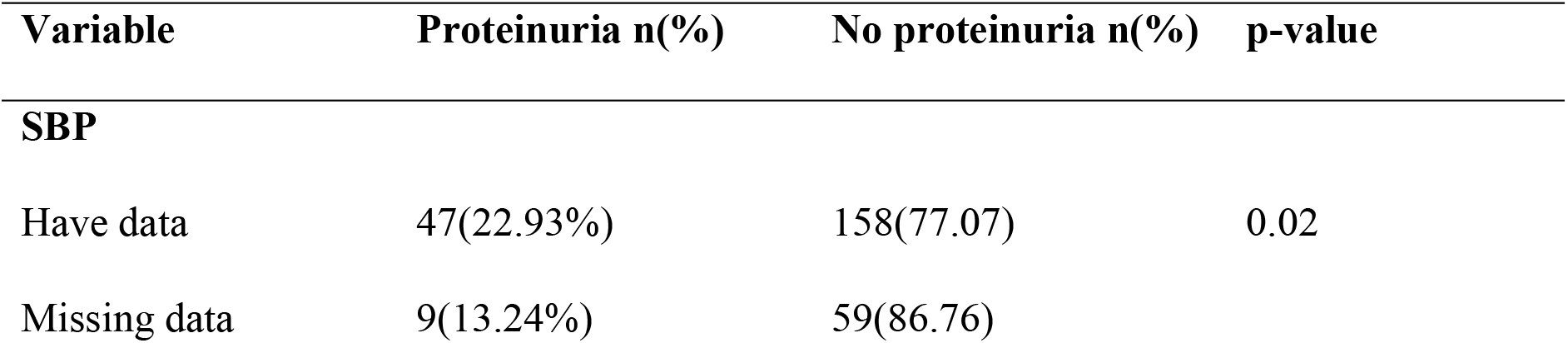
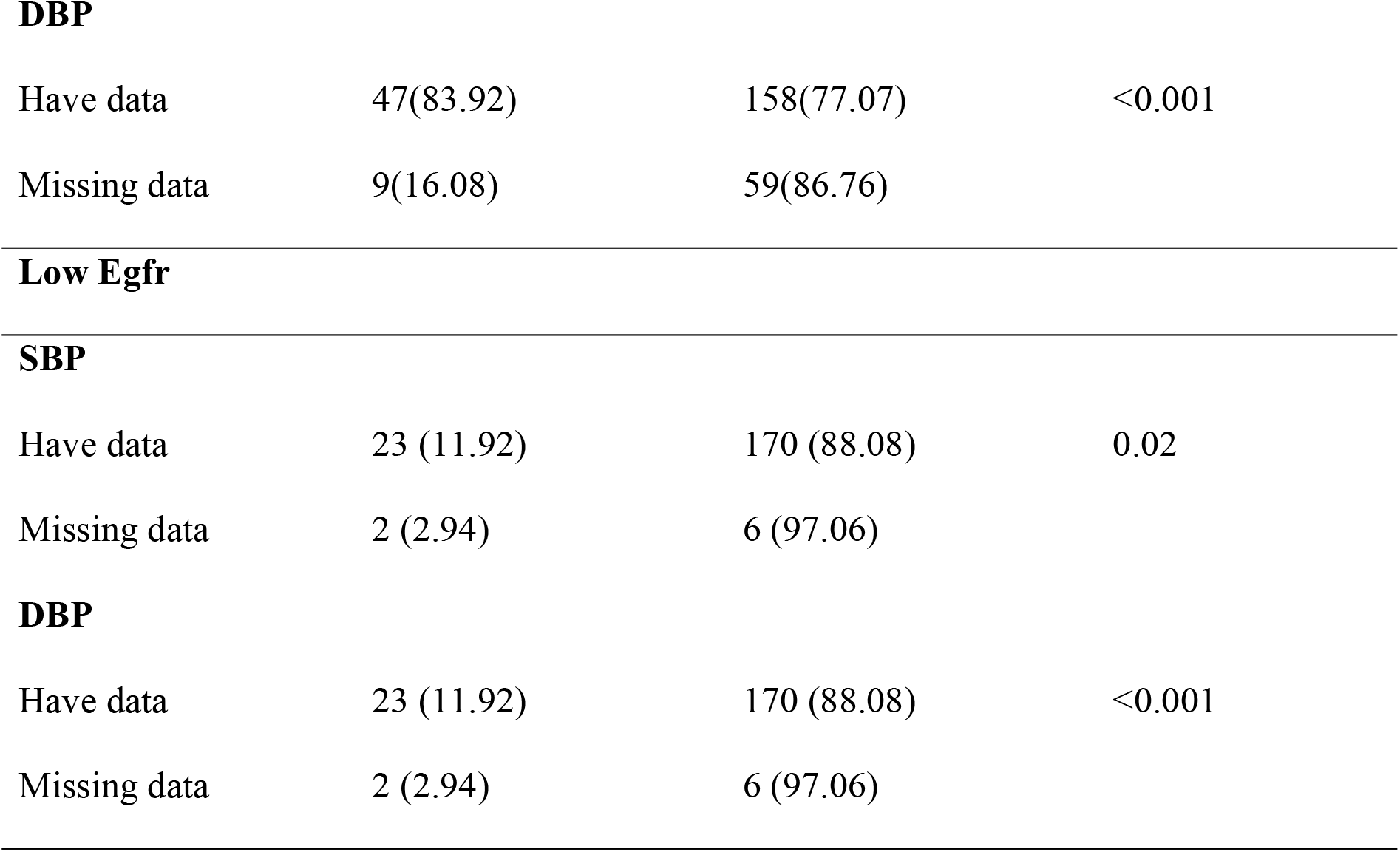
Comparison Between Participants with Missing Data and those With Proteinuria.

On bivariate analysis, the independent variables that were significantly associated with renal disease (low eGFR) were; SBP (OR= 2.6, p value = 0.19), DBP (OR = 43.66, p value = <0.001), Viral load (OR= 3.8, p value = 0.002) and ART Duration (OR =2.39, p value = 0.13). Results of bivariate analyses of factors associated with renal disease are summarised in table 5.

After adjusting for other covariates, factors significantly associated with renal disease (proteinuria) among the 261 participants that were recruited in February, 2020 to August, 2020 were; age (OR = 11.8, p-value = 0.001), WHO clinical stage IV of the disease (OR = 4.4, p value = 0.035), diastolic blood pressure (OR = 6.4, p value = 0.006) and viral load (OR = 7.0, p value <0.001). There was neither interaction nor confounding with any of significant variables.

In the above table, WHO disease stage 1 does not appear in the above table because there were no patients under this stage manifesting with proteinuria

After adjusting for other covariates, the only factor significantly associated with renal disease (low eGFR) among the 261 participants that were recruited in February, 2020 to August, 2020 was diastolic blood pressure. There was neither interaction nor confounding with any of the significant variables.

Prevalence of renal disease manifesting as proteinuria was statistically different from that of children with missing information and those with all information.

The prevalence of renal disease manifesting as low Egfr was statistically different from that of children with missing information and those with all information.

## Discussion

During the study period, the prevalence of proteinuria and low eGFR among children on ART attending the JCRC Pediatrics clinic was at 21.5% (95% CI; 17.0 – 27.0) and 9.6% (95% 7.0 – 14.0), respectively.

Proportions of renal disease were higher among children who had been on ART for more than 5 years as compared to those who had spent less than 5 years, those with a high viral load and high DBP also had higher proportions of proteinuria and low eGFR as compared to their respective categories.

The prevalences of proteinuria in this study was higher than that observed among children who attended the ART Clinic in Muhimbili National Hospital which were 7.1% and 5.8% was for low eGFR (Fredrick, Francis et al. 2016), this was lower than the prevalence of a reduced eGFR in this study and in Zimbabwe where proteinuria was 5%, lower than that in this study and low eGFR, 34.6%, higher than what was observed in this study. (Dondo, Mujuru et al. 2013). The prevalence of low eGFR was lower in this study as compared to that of 34.6% conducted in Zimbabwe (Dondo, Mujuru et al. 2013) but this was before the implementation of the test and treat policy, in which children presented with HIV - Associated Nephropathy, HIVAN, which also manifests as a low eGFR.

Differences in the criterion for defining renal disease are likely to contribute to the differences in prevalence. In this study, renal disease was defined as either proteinuria (+) on urine dipstick or eGFR below, 59ml/min1.73m2, this is similar to the Zimbabwean study (Dondo, Mujuru et al. 2013) which also considered presence of one of the two, to determine presence of renal disease. In contrast, the Nigerian study (Esezobor, Iroha et al. 2010) where eGFR derived from the cystatin C-based formula which is considered to be more accurate than the commonly used serum creatinine-derived formula since cystatin c is minimally affected by non-glomerular factors like diet and lean mass, showed a prevalence of 5.1% (Iduoriyekemwen, Sadoh et al. 2013). This is likely to affect the accuracy of this study in estimating renal disease because it is serum creatinine formula which was used to estimate the eGFR.

Most of the children were on TDF based regimen (29.9%)which could be responsible for the observed prevalence of renal disease as this regimen has been associated with renal disease (Campbell, Ibrahim et al. 2009) When only eGFR is used to determine renal disease, it does not give the true prevalence due to the fact that proteinuria is a marker of renal disease and can be present with normal eGFR (Iduoriyekemwen, Sadoh et al. 2013). This therefore justifies the use of either low eGFR or proteinuria to determine presence or absence of renal disease in this study.

According to a systematic review (Assaram, Magula et al. 2017), the prevalence of renal abnormalities with reference to proteinuria was high but had poor sensitivity and specificity to detect renal disease. This could be the reason for the high prevalence of proteinuria at 21.5% in this study.

The high prevalence of renal disease in this study may also be attributable to the assumption that renal disease majorly affects adults and the fact that priority to test renal functionality is only given to those who present with signs and symptoms before testing, this leaves most of the cases undiagnosed since renal function testing is not routinely done.

It is also notable that all the participants diagnosed with renal disease in this study were asymptomatic at the time of enrolment.

A high viral load was significantly associated with proteinuria (R = 7.0 p value <0.001) participants with a high viral load were 7 times more likely to present with proteinuria than those with a low viral load, which is consistent with findings from a Nigerian study whose p value = 0.014 (Esezobor, Iroha et al. 2010) and other study findings (Szczech, Gange et al. 2002, Cheung, Wong et al. 2007), these indicated that the less the immunosuppression - manifesting as high viral load, the higher the chances of renal disease manifesting as proteinuria. This is likely to be influenced by poor adherence to ART, thereby leading to elevation of the viral load.

Advanced stage of the disease (stage 4) was associated with proteinuria (0R=4.4 p value=0.035), implying that participants with advanced stage of the disease were 4.4 times more likely to manifest with proteinuria than those lower disease stage. This is consistent with findings from Nigeria where proteinuria frequently manifested among those with advanced disease stage (Esezobor, Iroha et al. 2010).

Diastolic BP was associated with proteinuria (OR = 6.5 p value = 0.005), implying that participants with a high Diastolic BP were 6.5 times more likely to present with proteinuria than those with normal BP, this is consistent with literature from other studies which indicated that diastolic hypertension is paralleled to the degree of renal disease, manifested as proteinuria (King 1962). The detrimental effect of blood pressure elevation might be difficult to establish because kidney disease itself elevates blood pressure (Hsu, McCulloch et al. 2005) Renal disease in patients may also be due to underlying focal segmental glomerulosclerosis (Kaplan 2010) as was commonly seen in the pre-ART era.

Diastolic blood pressure was the only covariate significantly associated with low eGFR (OR = 40.89 p-value < 0.001). Implying that participants with a high diastolic BP were 40.89 times more likely to present with reduced eGFR than those with a normal one. The prevalence of renal disease observed in this study indicates that in the nearby future, renal disease is likely to pose a significant financial burden on the health sector since all the participants were asymptomatic at the time of the study. This implies that at the time laboratory and clinical diagnoses of the disease are made, the disease may have progressed to end stage renal disease which is usually more complex and expensive to manage.

## Methodological considerations

The study population consisted of patients from different parts of the central and Eastern regions which comprise of people with characteristics generalizable to other parts of the country, as well as low-income countries of Sub Saharan Africa.

In this study, eGFR estimation using serum creatinine was used, serum creatinine assay is minimally affected by lean mass and diet, thereby affecting sensitivity of the method. Despite this, this method has been shown to be inexpensive and readily accessible as an indicator of renal disease.

## Conclusion

The factors that were significantly associated with renal disease manifesting as proteinuria include; age, high Diastolic BP and advanced clinical stage of the disease. It is also worth noting that at bivariate level of analysis, TDF based regimen was significantly associated with proteinuria.

A high Diastolic BP was significantly associated with renal disease, manifesting as low eGFR.

The prevalence of renal disease in this group of HIV-infected children on ART appeared to be attributable to age of the participant as children above 9 years were more likely to present with disease, high viral load, high diastolic bp and those with advanced stage of the disease.

The risk of chronic kidney diseases in this large contemporary cohort of HIV-infected individuals appeared to be attributable to a combination of HIV-related risk factors. In addition to the traditional risk factors cited in the literature, both, regimens containing tenofovir and HIV disease severity seem to be associated with kidney diseases. Assessment of proteinuria constitutes a novel method for chronic kidney disease staging in HIV-infected individuals and may be effectively used to stratify the risk of progression to end-stage renal disease.

## Recommendation

In light to the findings, assessment of proteinuria can be an effective, yet an affordable screening test for renal disease in order to enable earlier detection of disease, thereby lowering the risk of progression to end-stage renal disease. The National health guidelines on HIV management should consider conducting renal function monitoring every six months as is the case with viral load monitoring among this population. It is also important to closely monitor patients who are on TDF based regimen for renal functioning.

## Data Availability

All relevant data are within the manuscript and its Supporting Information files.

## Acknowledgements

I would like to thank the Research Office at Joint Clinical Research Center Lubowa, the laboratory staff and the study nurses that helped make this research process a success. I am also grateful to the participants whose patient files, blood and urine samples were used in data collection. I also extend my gratitude to my supervisors and mentors: Prof. Joan Kalyango, Dr. Bruhan Kaggwa, Assoc. Prof. Victor Musiime, Dr. Nicolette Nabukeera-Barungi and Assoc. Prof. Peter Waiswa for their precious time, knowledge and guidance.

## Author Contributions

**Conceptualization:** Bagoloire K. Lynn, Joan Kalyango, Kaggwa Bruhan, Henry Mugerwa, Nicolette Nabukeera Barungi and Victor Musiime

**Data curation:** Bagoloire K. Lynn and Sheillah Ansiima

**Formal analysis:** Bagoloire K. Lynn and Sheillah Ansiima

**Funding acquisition:** Bagoloire K. Lynn and Kaggwa Bruhan

**Investigation:** Bagoloire K. Lynn and Henry Mugerwa

**Methodology:** Bagoloire K. Lynn, Joan Kalyango and Kaggwa Bruhan

**Project administration:** Bagoloire K. Lynn and Henry Mugerwa

**Resources:** Bagoloire K. Lynn, Kaggwa Bruhan and Peter Waiswa

**Software:** Bagoloire K. Lynn and Sheillah Ansiima

**Supervision:** Bagoloire K. Lynn, Joan Kalyango, Henry Mugerwa, Nicolette Nabukeera Barungi and Victor Musiime

**Writing - original draft:** Bagoloire K. Lynn and Joan Kalyango

**Writing – review & editing:** Bagoloire K. Lynn, Joan Kalyango and Peter Waiswa

## References

Assaram, S., et al. (2017). “Renal manifestations of HIV during the antiretroviral era in South Africa: a systematic scoping review.” 6(1): 200.

Brennan, A., et al. (2011). “Relationship between renal dysfunction, nephrotoxicity and death among HIV adults on tenofovir.” 25(13): 1603.

Bygrave, H., et al. (2011). “Renal safety of a tenofovir-containing first line regimen: experience from an antiretroviral cohort in rural Lesotho.” 6(3): e17609.

Campbell, L., et al. (2009). “Spectrum of chronic kidney disease in HIV-infected patients.” 10(6): 329–336.

Cheung, C. Y., et al. (2007). “Prevalence of chronic kidney disease in Chinese HIV-infected patients.” 22(11): 3186–3190.

Dondo, V., et al. (2013). “Renal abnormalities among HIV-infected, antiretroviral naive children, Harare, Zimbabwe: a cross-sectional study.” 13(1): 75.

Dutta, A., et al. (2015). “The HIV treatment gap: estimates of the financial resources needed versus available for scale-up of antiretroviral therapy in 97 countries from 2015 to 2020.” 12(11): e1001907.

Ekrikpo, U. E., et al. (2018). “Chronic kidney disease in the global adult HIV-infected population: A systematic review and meta-analysis.” 13(4): e0195443.

Esezobor, C. I., et al. (2010). “Prevalence of proteinuria among HIV-infected children attending a tertiary hospital in Lagos, Nigeria.” 56(3): 187–190.

Foster, B. J. and M. B. J. T. A. j. o. c. n. Leonard (2004). “Measuring nutritional status in children with chronic kidney disease.” 80(4): 801–814.

Fredrick, F., et al. (2016). “Renal abnormalities among HIV infected children at Muhimbili National Hospital (MNH)—Dar es Salaam, Tanzania.” 17(1): 30.

Hsu, C.-y., et al. (2005). “Elevated blood pressure and risk of end-stage renal disease in subjects without baseline kidney disease.” 165(8): 923–928.

Iduoriyekemwen, N. J., et al. (2013). “Prevalence of renal disease in Nigerian children infected with the human immunodeficiency virus and on highly active anti-retroviral therapy.” 24(1): 172.

Kalyesubula, R., et al. (2014). “HIV-associated renal and genitourinary comorbidities in Africa.” 67: S68–S78.

Kaplan, N. M. (2010). Kaplan’s clinical hypertension, Lippincott Williams & Wilkins.

Kayange, N. M., et al. (2015). “Kidney disease among children in sub-Saharan Africa: systematic review.” 77(2): 272.

King, S. E. J. A. J. o. C. (1962). “Diastolic hypertension and chronic proteinuria.” 9(5): 669–674.

Kish, L. (1995). Survey Sampling. New York, NY: John Wily & Sons, Inc.

Msango, L., et al. (2011). “Renal dysfunction among HIV-infected patients starting antiretroviral therapy in Mwanza, Tanzania.” 25(11): 1421.

Onlin, O. (2013). “Perinatally HIV-infected adolescents.”

Ray, P. E., et al. (2004). “A 20-year history of childhood HIV-associated nephropathy.” 19(10): 1075–1092.

Swanepoel, C. R., et al. (2018). “Kidney disease in the setting of HIV infection: conclusions from a Kidney Disease: Improving Global Outcomes (KDIGO) Controversies Conference.” 93(3): 545–559.

Szczech, L. A., et al. (2002). “Predictors of proteinuria and renal failure among women with HIV infection.” 61(1): 195–202.

Trullàs, J.-C., et al. (2008). “Prevalence and clinical characteristics of HIV type 1-infected patients receiving dialysis in Spain: results of a Spanish survey in 2006: GESIDA 48/05 study.” 24(10): 1229–1235.

Uganda, G. o. (2016). “Uganda population-based HIV impact assessment.”

UNAIDS, G. A. J. G., Switzerland: World Health Organization Library (2016). “Global AIDS update 2016.”

